# Personalized Intracranial Circuit-Guided Deep Brain Stimulation for Treatment-Resistant Schizophrenia

**DOI:** 10.64898/2026.07.21.26358402

**Authors:** Sameer V. Rajesh, Ruchita M. Kumar, Clyde Knox, Oscar Araiza-Carranza, Jennifer Kriegel, Nader Pouratian, Carol A. Tamminga, Bradley C. Lega

## Abstract

We report a pilot study of deep brain stimulation (DBS) in three individuals with treatment refractory schizophrenia (TRS). DBS target selection was supported by an inpatient brain network mapping paradigm using intracranial electroencephalography. In conjunction with assessing stimulation-dependent symptom improvement, we derived spatiotemporally resolved profiles of psychosis and healthy brain states and identified stimulation targets that best shifted brain networks towards healthy states. Therapeutic stimulation sites were personalized for each participant but converged on salience network nodes including anterior cingulate and anterior insula. No significant adverse events were noted across participants. Moreover, two participants with chronic stimulation and clinical follow-up of at least 4 months reported significant improvement in both positive and negative symptoms, and treatment optimization is underway for the third. These critical pilot data establish the feasibility of personalized DBS guided by concurrent stimulation mapping, behavioral assessment, and biomarker monitoring as a treatment for TRS.

## Main

Schizophrenia is a debilitating psychiatric illness with an estimated prevalence between 0.25% and 0.5%. It is a leading cause of disability globally and is marked by high rates of treatment failure ^1–4^. Persistent functional disability and high relapse rates in treatment resistant schizophrenia (TRS), defined as insufficient response to effective treatments including antipsychotics and electroconvulsive therapy, contribute to enormous personal, social, and societal costs, with elevated suicide risk ^5^.

Deep brain stimulation (DBS) was developed for the treatment of movement disorders, but has been used to treat refractory psychiatric illness including treatment-resistant obsessive-compulsive disorder (trOCD) and treatment-resistant depression (TRD) ^6–10^. We posited that DBS may serve as a novel therapeutic option for TRS. As DBS seeks to modulate networks, psychiatric DBS has more recently been supported by the epilepsy surgery model. Putative networks underlying pathology are sampled across various symptom states using stereotactic electroencephalography (sEEG) to understand the effects of stimulation with high spatiotemporal resolution and optimize target choice ^11–15^. We applied this approach here.

Neuroimaging studies have implicated various brain regions in the pathophysiology of schizophrenia, guiding our implantation strategy. These regions comprise core components of canonical brain networks: (i) **salience network (SN)**: (dorsal anterior cingulate (dACC), anterior insula (aIns), (ii) **default mode network (DMN)**: medial prefrontal cortex (mPFC), posterior cingulate (PCC), and (iii) **central executive network (CEN)**: ACC, dorsolateral prefrontal cortex (dlPFC). To integrate these diverse findings, the “triple network model” posits that dysfunction within and among these networks may underlie pathological manifestations of schizophrenia ^16^. Additional studies describe pathological hyperactivation of **limbic** structures (amygdala, hippocampus), which may contribute to reduced emotional processing and memory-related cognitive deficits ^17–19^. The diverse set of potential therapeutic targets, the limited knowledge of precise neural features governing pathology within these networks, and potential heterogeneity across patient phenotypes necessitated a detailed mapping of both behavioral and electrophysiological response to stimulation across a wide array of targets within these networks in order to advance DBS as a therapy ^20^.

To facilitate brain state modeling across psychosis symptom states in a controlled, temporally precise fashion, we administered subanesthetic doses of ketamine (controlled by saline placebo) to induce transient psychosis as per previous protocols ^21–24^. We mapped psychosis states to underlying neural features and subsequently determined candidate therapeutic targets by assessing how stimulation altered network electrophysiology to resemble favorable (low symptom) or unfavorable (high symptom) psychosis states. We further narrowed target selection based on observed behavioral changes and participant interview responses as stimulation was being delivered.

We report the implementation, safety, and feasibility of this psychosis modeling and stimulation mapping paradigm in three individuals with TRS, along with long-term improvement in individuals with at least 4 months of clinical follow-up. Following the inpatient mapping phase, the first participant received DBS implantation in left dACC and right subgenual cingulate (sgCC)/medial orbitofrontal cortex (mOFC), the second in bilateral dACC, and the third in bilateral anterior insula (Table 1; Fig 2D).

**Table 1:** Clinical characteristics of study participants. PANSS: Positive and Negative Symptom Scale; A/V Auditory, Visual;

| Participant | Age Range/Sex | Primary Symptoms | Baseline PANSS | Adverse Events | DBS Targets |
| --- | --- | --- | --- | --- | --- |
| 1 | 31-40/M | A/V.<br>Hallucination | 90 | Surgical site pain;<br>Difficulty<br>sleeping | L: dACC<br>R: sgCC/mOFC |
| 2 | 41-50/M | Paranoia, Perse-<br>cutory/Religious<br>Delusion, A.<br>Hallucination | 103 | Loosening of one<br>sEEG bolt<br>necessitating<br>bedside removal | L: dACC<br>R: dACC |
| 3 | 21-30/M | Paranoia, A/V.<br>Hallucinations | 84 | Jaw pain | L: aINS<br>R: aINS |

These individuals were recruited based on established symptom intractability (persistence after an adequate trial of clozapine and/or ECT) and completed inpatient testing (mean: 8.67 days) followed by implantation of DBS electrodes into brain regions selected on the basis of sEEG mapping without serious adverse events across participants. Table 1 summarizes their clinical characteristics. Figure 1A shows anatomical locations of sEEG electrodes colored by region of interest or membership in a major brain network (SN, CEN, DMN, Limbic System). Figure 1B shows symptom severity measures over the outpatient optimization phase. Figure 1C is a schematic of the mapping paradigm.

**Figure 1:**
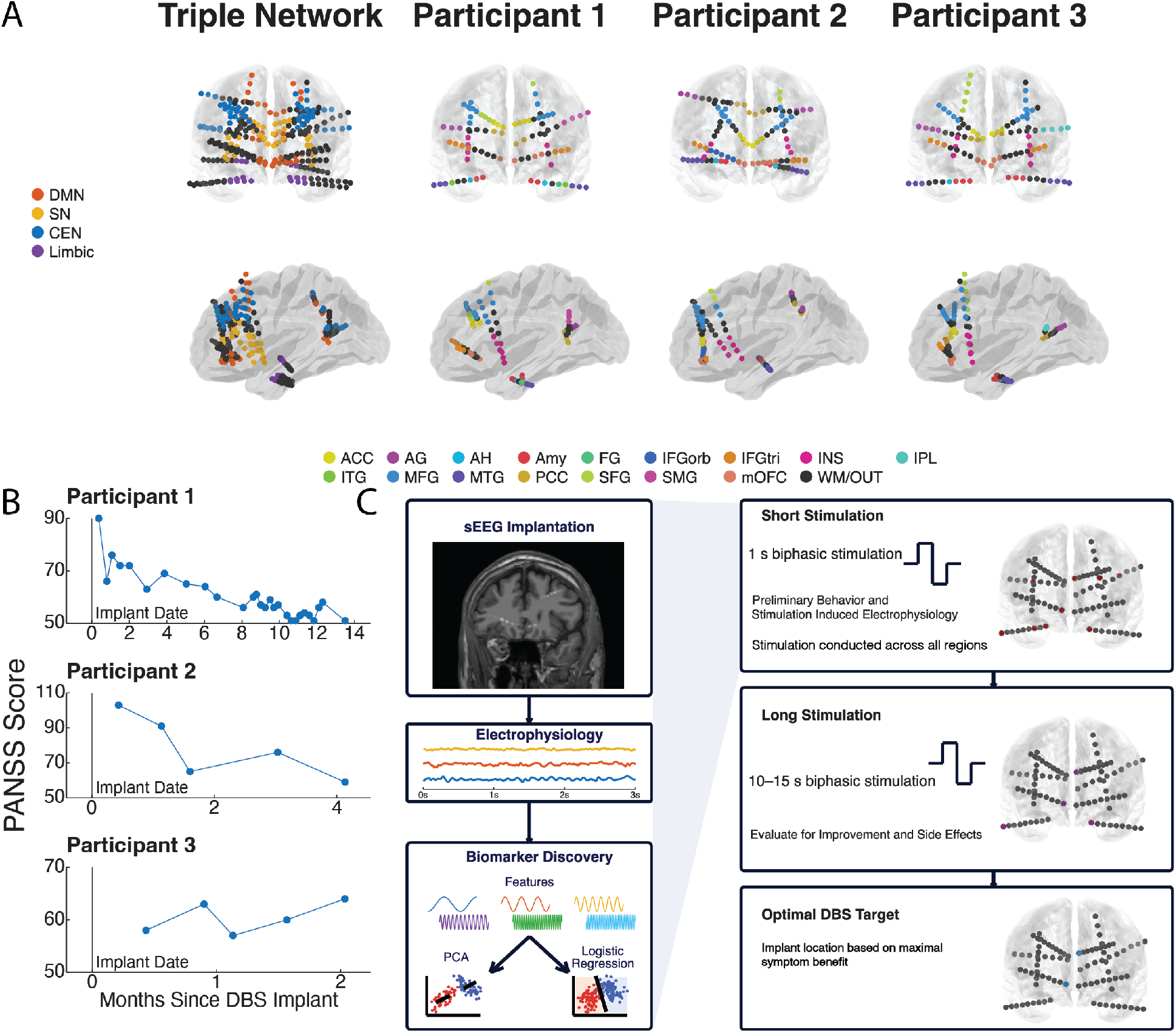
**A**. sEEG contact locations colored by network membership or region of interest. A/PCC: Anterior/Posterior Cingulate Cortex; AG: Angular Gyrus; AH: Anterior Hippocampus; Amy: Amygdala; FG: Fusiform Gyrus; IFGorb/tri: Inferior Frontal Gyrus, pars orbital/triangularis; INS: Insula; IPL: Inferior Parietal Lobule; I/MTG: Inferior/Middle Temporal Gyrus; M/SFG: Middle/Superior Frontal Gyrus; SMG: Supramarginal Gyrus; mOFC: medial orbitofrontal cortex; WM/OUT: White Matter, Extracranial. **B**. PANSS scores since DBS implantation. **C**. Schematic of mapping paradigm.

To decode the electrophysiologic patterns underlying disease states, we measured spatiospectral properties of intracranial signals contrasting each person’s most and least symptomatic recording periods using ketamine-induced or natural symptom variation. Ketamine elicited differential effects: participant 1 exhibited marked reduction of symptoms following drug administration, while participant 2 had ratable worsening of his symptoms. In participant 3, the impact of ketamine was equivocal but he experienced a spontaneous episode of more severe psychosis on hospital day 5 which we contrasted with a separate recording period (day 3) in which symptoms were mild.

Principal component analysis (PCA) reduced the dimensionality of the *∼* 300-dimensional (electrodes x frequency bands) feature space for each participant; a “principal improvement axis” (PIA) was defined as the vector pointing from centroid of the high symptom data cluster to the centroid of the low symptom data cluster, embedded in low dimensional space. Back-projecting this axis using PCA loading weights onto the original feature space revealed the participant-specific changes in regional oscillatory activity subserving symptom improvement (Fig. 2A); these were consistent with changes in electrode-averaged power spectral densities (Fig. 2B). Across participants we consistently observed suppression of limbic high frequency activity (HFA: gamma, high gamma) relative to other brain regions in low symptom states (Fig. 2A, 2C, S1). PCA decomposition also helped visualize the electrophysiological effect of ketamine, with uniformly reduced low frequency and increased high frequency power, consistent with previous findings and the putative mechanism of action (Fig. S1) ^25^.

**Figure 2:**
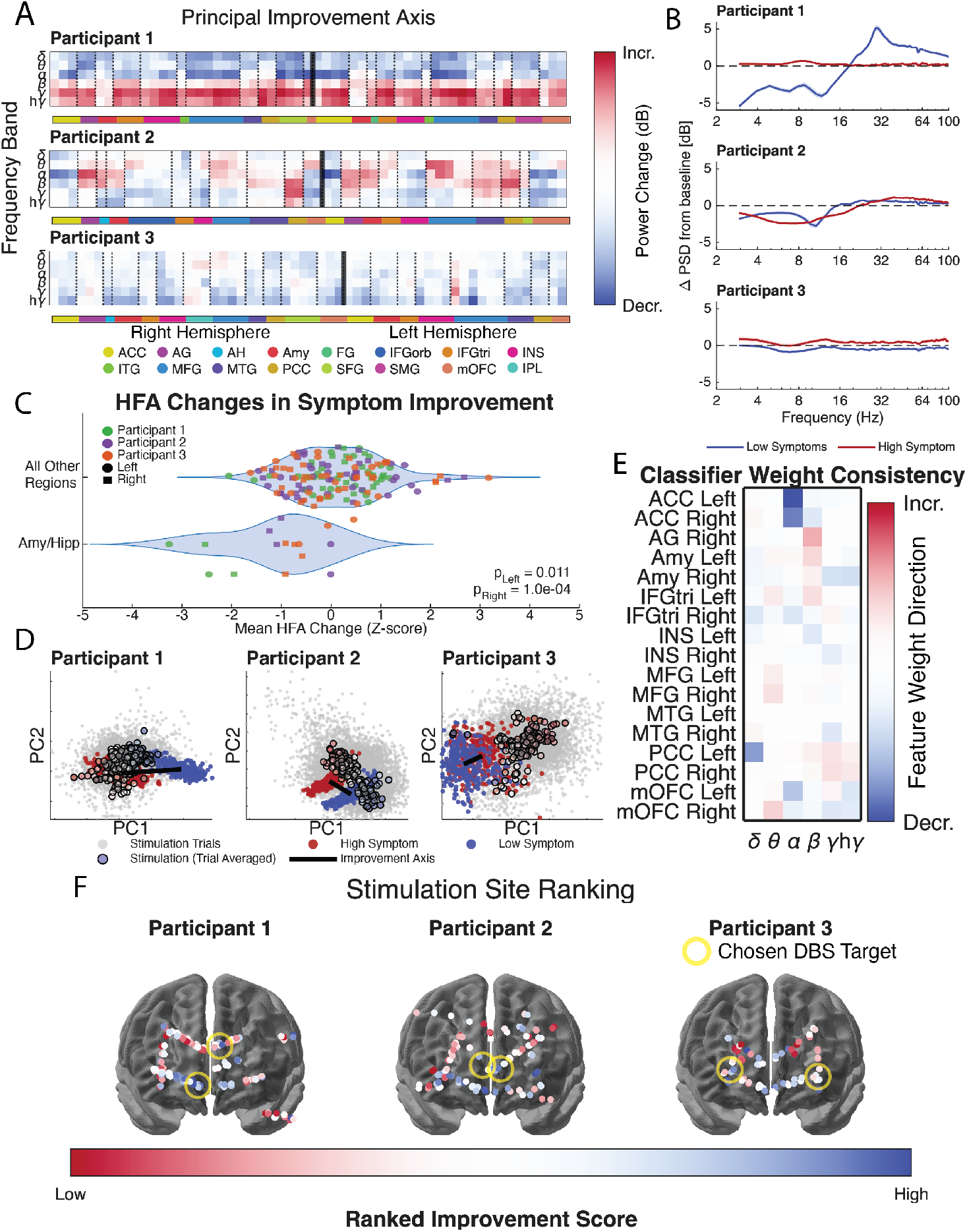
**A**. Regional oscillatory power changes underlying symptom improvement. **B**. Baseline-subtracted power spectral densities in high (red) and low (blue) symptom state (mean across channels excluding amyg-dala, hippocampus, 1 s.d. shaded). **C**. HFA of limbic electrodes is reduced in favorable symptom states relative to unfavorable ones. Statistics indicate permutation testing results.**D**. Scatter plot of spatiospectral feature vector from 1-second epochs from high/low symptom recordings using PC1/2. Gray points represent single trial post-stimulation spatiospectral feature vectors; **outlined** points represent trial-averages for each contact/stimulation parameter combination, colored by **improvement score rank** from most (blue) to least (red) improvement. Two-dimensional projection of PIA plotted in black. **E**. Consistency map of classifier weights across participants. Blue/Red: features that consistently decrease/increase across patients. **F**. Stimulation locations colored by stimulation improvement score. Only high frequency (100-120 Hz) stimulation experiments ranked.

We then computed baseline-normalized spatiospectral changes elicited by stimulation across implanted contacts. We projected the post-stimulation effect onto the PIA and scored the resultant position based on proximity to the low symptom state, with highest rank (highest score) indicating highest greater proximity (Figure 2D). To study convergent features across this cohort, we trained logistic regression classifiers (see *Methods*) to identify limited feature sets holding the most explanatory power in distinguishing psychotic from non-psychotic states, and measured similarity of these feature sets across individuals; this analysis revealed reductions in limbic HFA as well as alpha power in the ACC/mOFC to be associated with improvement (Fig. 2E).

We overlaid the stimulation sites colored by improvement score rank on the brain (Figure 2F). Stimulation of the amygdala and anterior hippocampus consistently worsened psychiatric symptoms across participants and were excluded from analysis due to early stopping of those trials. Stimulation of dACC/sgCC/mOFC elicited consistent greatest improvement in the first two participants (blue); stimulation of the aIns, and secondarily the sgCC/mOFC, elicited consistent improvement in the third. Final DBS implant locations were further refined based on observed improvement in symptoms elicited by stimulation (Fig 2F, Table 1).

This target selection approach appears to predict durable symtpom improvement. Participant 1 underwent extended outpatient therapy optimization with follow-up to determine clinical benefit, with a 1-year 43% PANSS score reduction from baseline, without suicidal ideation, psychiatric hospitalization, or wors-ened symptoms (Fig 1B). Importantly, the 4-week blinded discontinuation was completed with no event or symptom change. Participant 2 has undergone four months of open label stimulation optimization. Based on initial improvements that waned, and by analogy to stimulation approaches in other conditions, we initiated an hourly cycling stimulation schedule. This has led to significant (43%) reduction in PANSS score (Fig 1B). Both he and his family endorse strong symptom reduction. Participant 3 has experienced only transient benefits from stimulation modifications over 8 weeks. Adjustment and titration remain ongoing without adverse events.

We thus report the first use of personalized DBS for TRS in which target selection was guided by intracranial electrophysiology, behavioral assessment, and acute stimulation responses. Our findings provide initial evidence that acute stimulation can rapidly and reproducibly ameliorate psychosis symptoms and that combined neural and behavioral measures can identify candidate therapeutic targets. Our paradigm facilitates testing across broad cortical networks, which is particularly advantageous in TRS where optimal DBS target sites are unknown. Still, sEEG implantation is necessarily hypothesis driven, and our implantation array emphasized cortical and subcortical regions implicated in the triple network model. That therapeutic sites fall within the cingulo-insular salience network suggests that, despite inter–subject differences in the most favorable location, targeting within this circuit may offer a therapeutic strategy. Evaluation of additional patients may identify other favorable targets. Substantia nigra and nucleus accumbens are being pursued as DBS targets ^26,27^; future work may suggest a common therapeutic framework incorporating these regions as in OCD ^28^. Our biomarker findings are consistent with literature linking amygdalar and hippocampal activity with psychosis severity, although stimulation to these regions worsened symptoms ^18,19^. Further experience with direct brain recordings in TRS may identify physiologically defined disease phenotypes to further refine target selection.

There are limitations to our findings. Our initial reports are limited by small sample size and a selected study population We currently exclude individuals whose thought disorder precludes informed consent, and we require sufficient family support to facilitate clinic visits for stimulation testing. The cohort therefore represents highly motivated and supported individuals with TRS, albeit with severe symptoms. Broader enrollment will require validating our approach. Brain state mapping requires sufficient symptom variation to link acute changes in symptoms state with neural features, which may not occur spontaneously during intracranial mapping. Ketamine generated useful symptom variation, but generates a potential confound because it alters electrophysiologal signals. Nevertheless, symptom-related neural features converged across ketamine-induced and endogenous variation, supporting the validity of the pharmacological manipulation.. As data accumulate, a Bayesian framework integrating symptom-related features across participants may reduce or eliminate the need for pharmacological provocation. A further limitation of an inpatient mapping paradigm is that acute responses may not predict the chronic circuit effects or clinical benefit of DBS. Targets producing immediate symptom improvement may fail to confer durable benefit, whereas targets with meaningful chronic effects may show little acute response and therefore be overlooked. Correlating acute neural features with long-term outcomes is an important goal as our cohort expands.

Despite these limitations, the sustained improvements and overall safety observed in these initial participants are encouraging and provide support for personalized DBS in TRS. These findings warrant evaluation in larger trials designed to assess efficacy and durability.

## Data Availability

De-identified data produced in the present study are available upon reasonable request to the authors.

## Methods

### Participant Recruitment and Clinical History

Three male participants were recruited on the basis of established symptom intractability based on failure of medication treatment and electro-convulsive therapy. Participants with considerable thought disorder who could not plausibly give informed consent were not included. Participants were consented and the study was monitored as per UTSW IRB STU2023-0110 and trial information outlined in NCT06257056.

### Surgical Procedure and EMU stay

All participants were surgically implanted with 10 depth stereotactic electroencephalography (sEEG) electrodes (2 mm stainless steel contacts with 5 mm spacing; Depthalon, AdTech Corporation) spanning frontal, temporal, and parietal cortex, insula, cingulate, and mesial temporal lobe. Following implantation and post-anesthesia recovery, participants were admitted to the epilepsy monitoring unit for 8-9 days of inpatient monitoring and research study. Upon conclusion of the research aspects of the study and therapeutic site identification, participants were returned to the operating room, the depth sEEG electrodes were explanted, and DBS leads (Abbott Infinity 7 6662) were implanted into mapping-defined sites. The implanted pulse generator was also placed in this procedure.

### Intracranial Data Acquisition and Pre-processing

Intracranial electro-encephalography was acquired at 2000 Hz using a Nihon Koden system. We noted and excluded windows of decreased arousal post-operatively due to anesthesia. All signals were bipolar referenced against the adjacent contact on their respective probe. Bipolar signals where either contact was extracranial or both contacts were localized to white matter were ignored, and remaining contacts were assigned to a gray matter region based on the Automated Anatomical Labeling (AAL) scheme; localizations were reviewed by authors. The first participant retained 57 bipolar signals for analysis, the second 55, and the third 58. Data were then notch-filtered using 4th order Butterworth filters at 60 Hz, 120 Hz, 180 Hz, and 240 Hz to remove electrical line noise and subsequently down sampled to 500 Hz. These computations were supported by the EEGToolbox (Penn Computational Memory Lab). To extract band-limited spectral power, data were filtered using zero phase 4th order Butterworth filters (MATLAB *filtfilt*) into the following frequency bands: delta (2-4 Hz), theta (4-8 Hz), alpha (8-16 Hz), beta (16-32 Hz), gamma (32 - 55 Hz), and high gamma (65-100 Hz). Signal amplitude for each band was estimated as the square of the real part of the Hilbert transform (MATLAB *hilbert*)of the signal. A buffer of 500ms was applied to the ends of all signal prior to filtering and removed following amplitude calculation to eliminate edge artifacts. Baseline normalization was then applied using a decibel scale (see below sections for details on baseline windows). We further generated baseline normalized power spectral densities using Welch’s method with 50% overlapping 1-second windows (MATLAB *pwelch*)

### Modulation of Psychiatric Symptoms via Ketamine Administration

Ketamine was used to induce changes in psychiatric symptoms in absence of autonomous symptom development. During the 4 mornings following surgery, ketamine or placebo injections were administered intravenously under supervision of a clinician psychiatrist and a clinician neuroanesthesiologist. During ketamine sessions, the participant received an intravenous bolus of ketamine at 0.3 mg/kg (maximum total dose 60mg), administered by slow IV push over 1 minute. For placebo sessions, an equivalent volume of normal saline was administered using the same procedure. All sessions were separated by a minimum of 24 hours and ketamine and placebo session days were alternated, in a double-blind fashion. This modulation generated recordings with both high and low symptoms for each of the first two participants, while the third participant independently developed purely psychosis symptoms.

### Modeling of High and Low Symptom States

We compared data acquired during periods of greatest symptom intensity to those acquired during periods of least symptom intensity for each participant. For pharmacologically induced symptom paradigms, baseline activity was quantified from a 5-minute recording window beginning 10 minutes or prior to drug infusion onset. Analytical windows were selected as 5 minutes beginning 5 minutes after drug infusion was completed. In the third participant, we used a 5-minute long baseline window beginning 25 minutes prior to infusion, and a 5-minute long analytical window beginning 10 minutes prior to infusion to avoid inclusion of drug infusion data. Post-infusion power was normalized to baseline power on a logarithmic scale; for the third participant, the same baseline from the low symptom date was applied for normalization of both low and high symptom data, as we were interested in symptom variation and the participants high symptom day also had a symptomatic baseline.

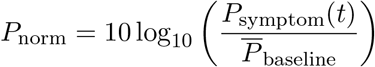

We averaged power data into non-overlapping 1 second epochs and treated these windows as individual samples for future analyses comparing symptom states.

### Stimulation Parameter Grid Search

Stimulation was delivered using the Blackrock Microsystems CereStim and Cerebus interface in a bipolar configuration. Biphasic pulse trains were administered every 5 seconds (1 second on, 4 seconds off) across a grid of amplitude and frequency combinations. Frequencies included 20, 50, 100, and 130 Hz; amplitudes ranged from 1–6 mA, with upper and lower bounds adjusted based on the presence of afterdischarges, side effects, and clinician recommendations at each stimulation site. Pulse width was kept constant at 150*µ*s. We followed established safety limits for brain stimulation set by the device manufacturer with a maximum allowed charge of 1.51 *µ*C delivered at the stimulation site.

For each stimulation trial, spectral power was quantified within a 1-second post-stimulation window beginning 1 second after completion of stimulation delivery and compared to a 500-ms pre-stimulation baseline window extending from 750 ms to 250 ms prior to stimulation onset. Data were preprocessed and normalized as described above, and power changes were expressed in decibels relative to baseline, yielding spatiospectral power estimates that could be directly compared to changes observed between high- and low-symptom states.

### Principal Component Analysis and Principal Improvement Axis Determination

To identify the primary transformation associated with symptom improvement across spatiospectral features, we constructed a low dimensional representation of the spectral power across regions (MATLAB *pca*). One-second long high and low symptom severity epochs were treated as separate observations, represented by feature vectors containing the concatenated channel-by-band power values across electrodes and bands. All epochs were combined into a single matrix and z-scored across observations on a feature-wise basis. Features with zero variance were retained by setting their standard deviation to one, and missing values were replaced with zero after normalization, such that missing measurements contributed no deviation from the population mean. PCA was then performed on the normalized data matrix. Principal components explaining at least 95% of the total variance were retained for subsequent analyses.

Within this PCA space, the centroid of the high and low symptom states (*µ*_high/low_) were computed as the mean of all data belonging to each condition. The principal improvement axis was defined as the unit vector connecting these two centroids:

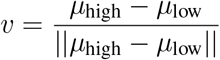

We then projected the improvement axis back into the full original feature space by multiplying the PCA loading matrix by *v*, producing a channel-by-frequency map representing the contribution of each spatiospectral feature to movement along the axis. Positive values for a specific spatiospectral feature indicated that movement along the axis towards the high symptom state was associated with increase in that feature, and vice versa.

After initial observations of potential attenuation of limbic HFA as symptoms improved, we attempted to quantify this result specifically. From the PCA maps projected back into the full orignal feature space, we extracted and z-scored the values for gamma and high gamma power change within frequency band across electrodes, then computed the median across bands for each electrode to get a summative, per-electrode, normalized measure of change in HFA with symptom improvement. We visualize these data points in violin plots in Figure 2D. We then assessed the statistical significance of the observed difference in limbic HFA using a patient and hemisphere stratified permutation test. Within each patient/hemisphere dataset, we calculated an observed delta between the mean HFA change across amygdalar/hippocampal electrodes and the mean across all other electrodes. We then shuffled electrode labels within each participant and hemisphere 10,000 times to generate a null distribution, and assessed the p value using as the fraction of shuffles where the permuted delta had an absolute value greater than the observed delta. We then averaged this p value across patients, which is acceptable as this is a Monte-Carlo permutation statistic. In this way we generated p values comparing electrodes within limbic regions to other regions within the same hemisphere.

### Therapeutic Site Determination

To quantify the electrophysiological impact of stimulation as movement along the principal improvement axis, pre-stim normalized post-stimulation data from each stimulation trial (which is the same dimension as the symptom state data due to being recorded from the same sites in 1 second epochs) was projected into the above described PCA space using the loading matrix *W* . Prior to projection the data was z scored using the mean *µ* and standard deviation *σ* of all data from the high/low symptom recordings:

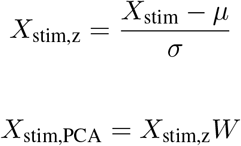

We then centered the data around the centroid of the high symptom cluster for each participant, and projected the resulting centered data point onto the improvement axis *v*. This value measured how far a stimulation trial lay along the improvement axis. Scores were normalized by the length of the improvement axis, so that a score of 0 corresponded to no movement, while a score of 1 corresponded to complete shift towards the low symptom state. Scores were then averaged across all trials at the same stimulation contact with the same stimulation parameters. Each contact/parameter combination was then separately ranked (within hemisphere) based on this improvement score. Brain plots in Figure 2F show ranking using only experiments conducted at high frequency stimulation.

### Logistic Regression Model Identification of Discriminative Features

We trained an elastic-net regularized logistic regression classifier to discriminate high- and low-symptom states using region-by-band activity. For each participant, power values from all electrode-frequency band combinations were concatenated into a feature matrix, with recordings from the high-symptom state assigned a label of 1 and recordings from the low-symptom state assigned a label of 0. Features containing non-finite values were excluded prior to model fitting.

Data were randomly partitioned into training (70%) and testing (30%) sets while preserving the relative number of samples from each symptom state. Feature standardization was performed using the mean and standard deviation of the training set, and the resulting normalization parameters were subsequently applied to the held-out test set. Elastic-net logistic regression models were fit using a range of mixing parameters (*α* = 0.1, 0.3, 0.5, 0.7, 0.9) and regularization strengths (*λ* = 10^*™*4^) *™* (10^0^), 27 logarithmically spaced values). Hyperparameters were selected by minimizing the negative log-likelihood on the held-out test data. The final model was then refit using the optimal hyperparameters. We did not apply cross-validation here and thus do not emphasize the classification accuracy, which was generally high. We maintain that the emphasis of this approach was as a secondary means of discovering the most discriminative spatiospectral features.

The resulting model coefficients were reshaped into electrode-by-frequency-band weight maps, yielding a quantitative measure of the contribution of each spatiospectral feature to discrimination between symptom states. Positive coefficients indicated features associated with the high-symptom state, whereas negative coefficients indicated features associated with the low-symptom state.

Cross participant similarity was assessed by first aggregating classifier weights within each region using the median across contacts to obtain region-resolved representations. Only ROIs shared across all participants were retained, and matrices were reordered to a shared order for direct comparison. A consistency score was computed at each feature–label element by combining signed agreement and magnitude consistency across all pairwise participant comparisons. The resulting values were averaged and normalized by their maximum absolute magnitude. The final matrix was visualized as a heatmap of bands versus shared labels, indexing reproducibility of classifier-derived weights across datasets.

### Open Label Phase

Following the parameter search, and utilizing the participant specific parameters determined, we conducted an eight month open label phase to systematically assess both the safety and efficacy of open loop stimulation for the treatment of schizophrenia. Our primary endpoint was lack of adverse events and successful completion of the therapy. Our pre–defined efficacy endpoint, and the study threshold for clinical significance, was *≥* 25% reduction in total PANSS score from presurgical baseline to the start of the blinded discontinuation period (8 months after implantation). We also implemented cycling protocols in the second participant, where DBS as switched on and off at regular intervals to provide a varying stimulation pattern that has demonstrated efficacy in other contexts.

Secondary outcomes tracked positive and negative PANSS sub-scales separately. Our hypothesis being that the participant would achieve clinical response and that improvements in positive symptoms would precede improvements in negative or cognitive symptoms. Safety was defined using validated clinical thresholds: *≤* 10% worsening in total PANSS score, *≤* 6 point increase in Calgary Depression Scale/MADRS, and *≤* 0.5 SD decline in MATRICS Consensus Cognitive Battery performance. Inpatient safety was monitored by way of postoperative imaging, structured neuro-psychiatric evaluations, and real-time side-effect screening during sEEG stimulation. And outpatient safety was tracked every two weeks with standardized metrics, medication adjustments, and structured feedback. Finally, safety was also assessed during a blinded discontinuation phase.

