## SupplementalFigure1 for "Personalized Intracranial Circuit-Guided Deep Brain Stimulation for Treatment-Resistant Schizophrenia"

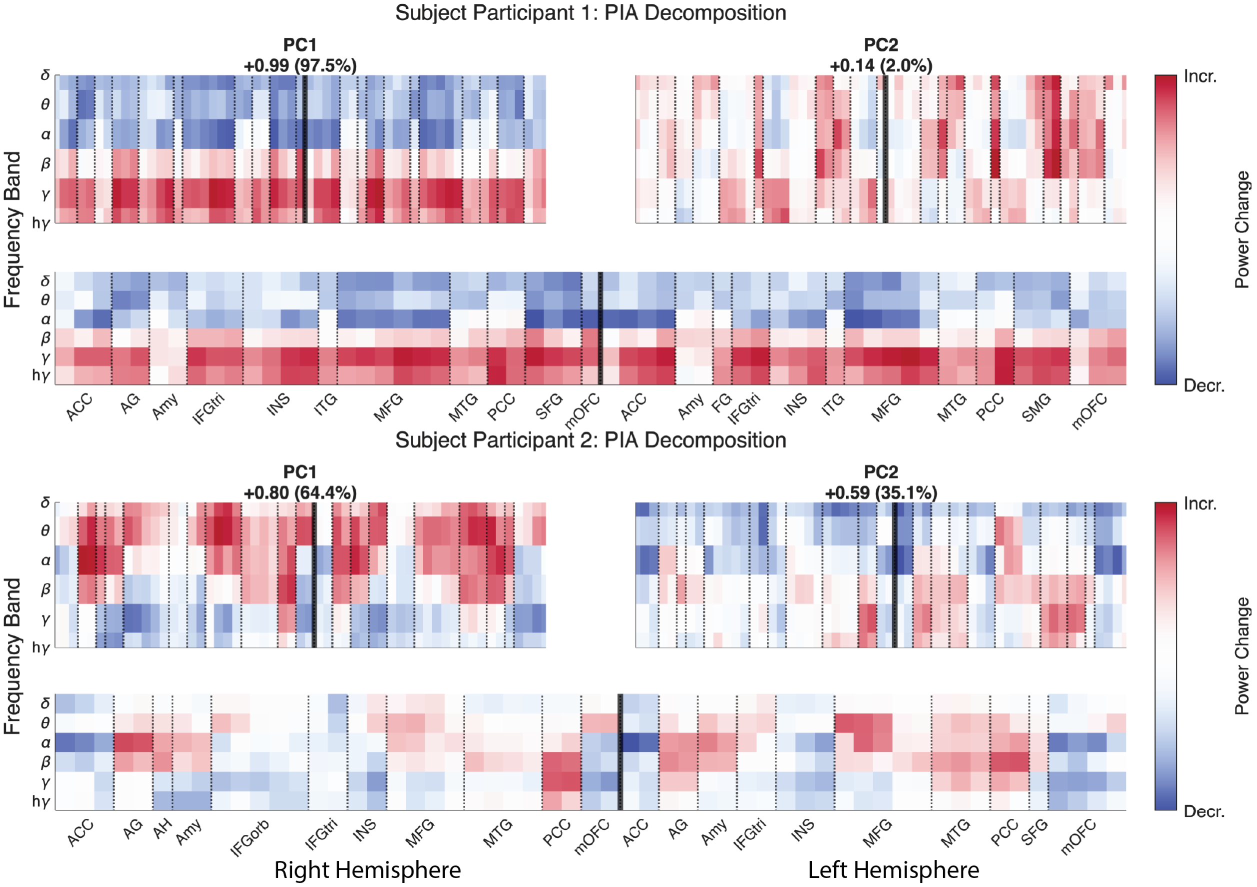


**Figure 1: Regional oscillatory power changes associated with ketamine** In Participants 1 and 2, ketamine was used to induce symptom variability. We show the top two PCs for these participants to decompose the change underlying symptom improvement. As ketamine improved the first participant's symptoms and worsened the seconds, the top row of figures thus shows the effect of ketamine relative to placebo while the second row shows the negative of that effect. We note the similarity between Participant 1 PC1 and the negative of Participant 2 PC1 as evidence of potentially convergent dominant electrophysiological effect of ketamine across Participants to suppress low frequencies and amplify high frequencies.
